# Effect of metabolic syndrome components on the risk of malignancy in patients with gallbladder lesions

**DOI:** 10.1101/2020.06.06.20123992

**Authors:** Zheng Deng, Yan Xuan, Xinxing Li, William J Crawford, Zhiqing Yuan, Zhoukan Chen, Anastasia Brooks, Xiaowen Liang, Haolu Wang, Tao Chen

**Author notes:** Haolu Wang and Tao Chen share co-corresponding authorship. Haolu Wang, Tao Chen. Zheng Deng, Yan Xuan and Xinxing Li contributed equally to this work.

## Abstract

**Background and Aims:** The malignant potential of gallbladder lesions is debated, and there is limited guidance on surveillance. Predicting their risk of malignancy could help clinicians manage and potentially improve prognosis. We evaluated the independent and joint effects of metabolic syndrome components on the risk of malignancy among patients with gallbladder lesions.

**Methods:** Using a multicenter database, consecutive patients with pathologically confirmed gallbladder lesions between 2012 and 2019 were identified. Univariate and multivariate logistic regression analyses were used to evaluate the effects of metabolic syndrome components (diabetes, hypertension, dyslipidemia, obesity) as additive or combined indicators for the risk of malignancy. Unadjusted and adjusted odds ratios were calculated.

**Results:** Of the 625 patients, 567 patients were identified with benign gallbladder lesions and 58 patients with gallbladder cancer (GBC). Among all metabolic syndrome components, the proportion of GBC patients with dyslipidemia (63.8%) was significantly greater than benign gallbladder polyps (42.2%, P = 0.002). In multivariate logistic regression analysis, dyslipidemia was associated with a 2.67-fold increase in the risk of GBC (95% confidence interval 1.17-6.09). Dyslipidemia is an independent risk factor for malignancy (adjusted odds ratio 2.164, 95% confidence interval 1.165-4.021), regardless of whether the other risk factors and metabolic syndrome components are combined. Dyslipidemia (adjusted odds ratio 2.164, 95% confidence interval 1.165-4.021) and decreased high-density lipoprotein (HDL, adjusted odds ratio 3.035, 95% confidence interval 1.645-5.600) were closely associated with increased risk of malignancy.

**Conclusions:** Dyslipidemia is associated with a 2.67-fold increase in the risk of malignancy, regardless of the presence of other metabolic syndrome components. Dyslipidemia is an independent risk factor for malignancy in patients with gallbladder lesions.

## Introduction

Gallbladder lesions have become more common now with the increased use of ultrasound[1]. The incidence rate of gallbladder lesions is approximately 5-9.9% of the population[2]. Cholesterol gallbladder polyps are the most common type of benign gallbladder polyps (BGP). Some BGP, such as adenomas, are considered with malignant potential [1]. About 3-8% of all gallbladder lesions are malignant[3]. Current European consensus guideline suggests that all gallbladder lesions greater than 10 mm should be surgically removed and smaller lesions between 6 and 9 mm with co-existing presence of high risk factors are deemed to justify cholecystectomy [4]. However, there is no clear guidance on how to manage patients who were not offered surgical treatment[5]. It has been suggested patients with gallbladder lesions less than 10 mm may be safely followed conservatively[1, 4]. For such surveillance to be cost-effective, better risk stratification is needed to guide targeted surveillance. Predicting malignancy by better understanding risk factors would also allow clinicians to more effectively plan secondary prevention efforts of gallbladder cancer (GBC) for patients with gallbladder lesions.

Some manifestations of metabolic syndrome (such as diabetes, hyperlipidemia and obesity) are risk factors for gallbladder lesions[6]. Several studies support diabetes and hyperlipidemia may also be associated with biliary tract cancers[7]. These metabolic syndrome components are also readily identifiable and potentially modifiable, rendering them as ideal targets not only for risk stratification but also for risk modification, which would be useful to both predicting prognosis and preventing complications.

Although previous studies suggest a potential association between metabolic syndrome components and the risk of GBC[8], there remains doubt about the strength and extent of this association. Most available cohort studies followed only a limited number of patients with gallbladder lesions with incomplete risk factor data[9, 10]. The frequent interaction and co-occurrence of these metabolic syndrome components further complicates the analysis of each component’s specific contribution to the risk of malignancy in patients with gallbladder lesions.

To fill this gap in the literature, we conducted a retrospective cohort study of 625 patients with pathologically confirmed gallbladder lesions from three hospitals in China. The independent and joint effects of syndrome components on the risk of malignancy were evaluate.

## Method

### Study population

Using a multicenter database, a total of 625 consecutive postoperative patients who pathologically diagnosed with gallbladder lesions from February 2012 and December 2019 at three Chinese hospitals (Renji Hospital, Ruijin Hospital and Shanghai Changzheng Hospital) were identified. A standardised data form was created to collect all relevant information such as age, gender, body mass index, hypertension, diabetes, the data of ultrasonography including number of gallbladder lesions (single or multiple), accompanying with stones and maximum diameter of lesions and laboratory findings including liver function tests and lipid profiles. The inclusion criteria for the patients with gallbladder lesions were as follows: (a) patients underwent surgical treatment and had pathologically confirmed diagnosis; (b) examination with ultrasonography was performed within 1 month before surgery. We excluded patients diagnosed with other cancers within the five years before the date of gallbladder lesions diagnosis to ensure the metastatic cancers were excluded. Patients who are missing any of the required data were excluded. Patients who were younger than 18 years are excluded. Patients who comorbid with bile duct abnormalities such as choledochal cyst and gallbladder abnormalities such as porcelain gallbladder were excluded. Dyslipidemia was defined when any of the following criteria were satisfied: 1) hypercholesterolemia: ≥ 6.2 mmol/L; 2) hypertriglyceridemia: ≥ 2.3 mmol/L (200 mg/dL); 3) low HDL cholesterol: <1.03 mmol/L (40 mg/dL) in men and <1.29 mmol/L (50 mg/dL) in women. Obese was identified by BMI ≥25kg m^-2^ according to the WHO classification for Asian populations.

### Laboratory tests

All study participants were subjected to overnight fasting after which blood samples were drawn by clinical nurses for laboratory examination. Fasting blood glucose, total cholesterol, triglyceride, high density lipoprotein (HDL).

### Statistical analysis

Data on continuous variables were expressed as mean ± standard deviation (SD), the categorical variables are summarized as frequencies and percentages. We compared the demographic features and some of prevalence of risk factors between patients with BGP and GBC. We examined the risk of gallbladder lesions associated with features of metabolic syndrome components, including hypertension, diabetes, obesity and dyslipidemia. T–tests were conducted for continuous variables and χ2 tests for categorical variables. Odds ratios (OR), 95% confidence interval (CI) and p values were calculated for each variable. The multivariate logistic regression models were constructed to examine risk of GBC compared BGP group. Then we constructed univariate and multivariate model to test the joint association between dyslipidemia and the other metabolic syndrome components and restricted one risk factor without the others. Models were adjusted for age and gender. The results were considered statistically significant when p values were < 0.05. T–tests and χ2 tests were used to judge the significance of each variable. The adjusted ORs and 95% CIs were calculated for each parameter estimate. Statistical analyses were performed by SPSS version 25 (IBM Co., Armonk, NY, USA).

The study was conducted in accordance with the Declaration of Helsinki. The study protocol was approved by the Shanghai Jiaotong University School of Medicine, Renji Hospital Ethics Committee (Ethical approval no. 2016-045).

## Results

### Patient characteristics

We identified 625 patients with gallbladder lesions. Of these patients 567 had BGP, while 58 had GBC (Table 1). There was no significant difference in gender between these two groups. Some previous researches showed that the results were consistent with our study[10], and some literature reported that gender is one of the factors[11]. The mean age of the patients in the two groups was 49.94 ± 12.73 and 65.84 ± 9.98 years old, respectively. Patients in GBC group were significantly older than patients in BGP (P< 0.001) as well as patients with GBC were older than patients with gallbladder adenomatous polyps (P< 0.001, Table S1). Multiple lesions were found more frequently in patients with BGP (64.6%). than in patients with GBC (30.3%, P < 0.001). The level of total bilirubin was notably higher in GBC group than BGP group (P = 0.001) and adenomatous polyps’ group (Table S1). Previous literature has reported that elevated bilirubin level or jaundice are the independent risk factors for GBC, which is consistent with the results of this study[11, 12]. The study revealed a significantly different mean lesion diameter of 6.49 ± 4.60 mm, 9.27 ± 5.70 mm and 26.26 ± 22.66 mm for the BGP, adenomatous polyps, and GBC groups respectively (all P < 0.001). Accompanying gallstones were more common in the GBC group (3.4%) than in the BGP group (3.4% vs 20.8%, P = 0.001) as well as between in GBC group and adenomatous polyps’ group (P = 0.001, Table S1).

**Table 1.** Baseline characteristics of patients with gallbladder lesions

|  | Benign gallbladder polyps<br>(n = 567) | Gallbladder cancer<br>(n = 58) | P value |
| --- | --- | --- | --- |
| Gender |  |  | 0.990 |
| Male | 284 (50.1) | 29 (50.0) |  |
| Female | 283 (49.9) | 29 (50.0) |  |
| Age, mean (SD) | 49.94 (12.73) | 65.84 (9.98) | **<0.001** |
| Number of polyps |  |  | **<0.001** |
| Single | 201 (35.4) | 41 (70.7) |  |
| Multiple | 366 (64.6) | 17 (29.3) |  |
| Total bilirubin | 18.40 (47.80) | 44.15 (88.99) | **0.001** |
| Maximum diameter of polyps | 6.49 (4.60) | 26.26 (22.66) | **<0.001** |
| With stones | 118 (20.8) | 2 (3.4) | **0.001** |
| BMI $\geq 25$ | 202 (35.6) | 13 (22.4) | **0.044** |
| Hypertension | 275 (48.5) | 34 (58.6) | 0.142 |
| Diabetes | 84 (11.3) | 10 (17.2) | 0.181 |
| Dyslipidemia | 239 (42.2) | 37 (63.8) | **0.002** |
| Total cholesterol $\geq 6.2$ | 87 (15.3) | 6 (10.3) | 0.308 |
| TG $\geq 2.3$ | 72 (12.7) | 9 (15.5) | 0.543 |
| Decreased HDL | 144 (25.4) | 31 (53.4) | **< 0.001** |

TG: Triglycerides; HDL, high-density lipoprotein. BMI: body mass index.

Obesity was more common in the GBC group compared to patients in the BGP group (35.6% vs 22.4%, P = 0.044). Dyslipidemia was more common in the GBC group than the BGP group (63.8% vs 42.2%, P = 0.002). Within this, decreased HDL was more common in the GBC group (53.4%) than the BGP group (25.4%) and adenomatous polyps’ group (P < 0.001, Table S1). There were no significant differences in the prevalence of raised total cholesterol (15.3% vs 10.3%) and triglycerides (TG, 12.7% vs 15.5%) between the two groups (Table 1). There was no difference in prevalence of hypertension (48.5% vs 58.6) and diabetes (11.3% vs 17.2%) between the two groups.

### Independent risk factors for GBC

Univariate and multivariable analyses were performed to identify independent risk factors for GBC (Table 2). A single factor regression analysis on all forecast indicators demonstrated that age, number of polyps, total bilirubin, maximum diameter of polyps, accompanying stones, BMI and dyslipidemia were associated with an increased risk of GBC. Dyslipidemia was associated with a 2.4-fold increase in the odds of GBC (P = 0.002).

**Table 2.** Univariate and multivariate analyses of the risk factors for gallbladder lesions

|  | Univariate analysis |  |  | Multivariate analysis |  |  |
| --- | --- | --- | --- | --- | --- | --- |
|  | OR | 95% CI | P | Adjusted OR | 95% CI | P |
| Age | 1.130 | 1.095-1.165 | **< 0.001** |  |  |  |
| Gender | 0.996 | 0.580-1.711 | 0.990 |  |  |  |
| Number of polyps | 0.228 | 0.126-0.411 | **< 0.001** | 1.307 | 0.566-3.020 | 0.531 |
| Total bilirubin | 1.005 | 1.001-1.008 | **0.005** | 1.003 | 0.998-1.009 | 0.222 |
| Maximum diameter of polyps | 1.180 | 1.133-1.230 | **< 0.001** | 1.162 | 1.107-1.219 | **< 0.001** |
| Stones | 0.136 | 0.033-0.565 | **0.006** | 0.178 | 0.036-0.882 | **0.034** |
| Hypertension | 1.504 | 0.870-2.602 | 0.144 | 1.116 | 0.478-2.609 | 0.799 |
| Diabetes | 1.637 | 0.790-3.395 | 0.185 | 0.768 | 0.240-2.465 | 0.768 |
| Dyslipidemia | 2.418 | 1.380-4.237 | **0.002** | 2.674 | 1.173-6.094 | **0.019** |
| BMI $\geq 25$ | 0.522 | 0.275-0.991 | **0.047** | 0.552 | 0.228-1.338 | 0.188 |

Model adjusted for age and sex.
OR, odds ratio; CI, confidence interval;

These risk factors were then included in the multivariate analysis, which was adjusted for age and gender. The odds of GBC patients being diagnosed with dyslipidemia still increased by 2.6-fold and size of polyps had modest effect with OR of 1.162. In another multivariate analysis, we found that decreased HDL was associated with a 5-fold increase in GBC risk (95%CI: 1.502-16.801, P=0.009, Table S2). The number of polyps, total bilirubin, diabetes and BMI were not associated with GBC in the multivariate analysis.

### Joint associations of metabolic syndrome components with GBC

Pathological metabolic syndrome components often coexist in patients. We therefore sought to determine whether specific combinations of them were associated with increased risk of progression to GBC, while ruling out interference from other metabolic factors. Table 3 summarises the different types of metabolic characteristics found in the two groups. Numerous combinations of metabolic conditions were more common in the GBC than the BGP, including diabetes without a BMI ≥ 25, dyslipidemia without a BMI≥25, hypertension without a BMI ≥ 25, diabetes without hypertension, both dyslipidemia and hypertension, and patients with both dyslipidemia and diabetes.

**Table 3.** Types of metabolic traits of patients with gallbladder polypoid lesions

| Metabolic syndrome components | Benign gallbladder polyps<br>(567) | Gallbladder cancer<br>(58) | P value |
| --- | --- | --- | --- |
| Hypertension | 275 (48.5) | 34 (58.6) | 0.142 |
| Hypertension Excluding Diabetes | 230 (40.6) | 29 (50.0) | 0.165 |
| Hypertension Excluding Dyslipidemia | 152 (26.8) | 10 (17.2) | 0.113 |
| Hypertension Excluding BMI ≥ 25 | 153 (27.0) | 24 (41.1) | **0.020** |
| Diabetes | 84 (11.3) | 10 (17.2) | 0.181 |
| Diabetes Excluding Hypertension | 19 (3.4) | 5 (8.6) | **0.047** |
| Diabetes Excluding BMI ≥ 25 | 35 (6.2) | 7 (12.1) | 0.088 |
| Diabetes Excluding Dyslipidemia | 38 (6.7) | 2 (3.4) | 0.335 |
| Dyslipidemia | 239 (42.2) | 37 (63.8) | **0.002** |
| Dyslipidemia Excluding Hypertension | 116 (20.5) | 13 (22.4) | 0.726 |
| Dyslipidemia Excluding Diabetes | 213 (37.6) | 29 (50.0) | 0.064 |
| Dyslipidemia Excluding BMI ≥ 25 | 147 (25.9) | 29 (50.0) | **< 0.001** |
| Dyslipidemia and Hypertension | 123 (21.7) | 24 (41.4) | **0.001** |
| Dyslipidemia and Diabetes | 26 (4.6) | 8 (13.8) | **0.003** |
| Dyslipidemia and BMI ≥ 25 | 92 (16.2) | 8 (13.8) | 0.630 |
| BMI ≥ 25 | 202 (35.6) | 13 (22.4) | **0.044** |

Pathological metabolic syndrome components often coexist in patients. We therefore sought to determine whether specific combinations of them were associated with increased risk of progression to GBC, while ruling out interference from other metabolic factors. Table 3 summarises the different types of metabolic characteristics found in the two groups. Numerous combinations of metabolic conditions were more common in the GBC than the BGP, including diabetes without a BMI ≥ 25, dyslipidemia without a BMI≥25, hypertension without a BMI ≥ 25, diabetes without hypertension, both dyslipidemia and hypertension, and patients with both dyslipidemia and diabetes.

To verify whether these factors increased GBC risk, both univariate and multivariate analyses were performed (Table 4). In the univariate analysis, we found that hypertension without BMI ≥ 25 (OR=1.910, 95% CI:1.097-3.325), with dyslipidemia without BMI ≥ 25 (OR=2.857, 95% CI: 1.652-4.924), with both dyslipidemia and hypertension (OR=2.548, 95% CI: 1.456-4.458) and with both dyslipidemia and diabetes (OR=3.329, 95% CI: 1.432-7.740) were associated with increased odds of GBC. The multivariate analysis model was adjusted for age and gender, and dyslipidemia excluding diabetes group and dyslipidemia excluding obese group were significantly associated with GBC. Dyslipidemia without diabetes, and dyslipidemia without BMI≥25 were both associated with an approximately 1.9- and 2.5-fold increase in GBC risk respectively. The trait of hypertension without dyslipidemia seem to be a protective factor for patients (OR=0.438, 95% CI: 0.201-0.953). There were no other significant interactions among dyslipidemia and other risk factors. The results indicated that regardless of whether other risk factors are combined, dyslipidemia is indeed an independent risk factor for increasing the odds of GBC.

**Table 4.** Univariate and multivariate analyses of the association of types of metabolic syndrome components for gallbladder polypoid lesions

|  | Univariate analysis |  |  | Multivariate analysis |  |  |
| --- | --- | --- | --- | --- | --- | --- |
|  | OR | 95% CI | P | Adjusted OR | 95% CI | P |
| Age | 1.130 | 1.095-1.165 | **< 0.001** |  |  |  |
| Gender | 0.996 | 0.580-1.711 | 0.990 |  |  |  |
| Hypertension | 1.504 | 0.870-2.602 | 0.144 | 0.911 | 0.496-1.675 | 0.765 |
| Hypertension Excluding Diabetes | 1.465 | 0.853-2.518 | 0.167 | 1.221 | 0.671-2.222 | 0.512 |
| Hypertension Excluding Dyslipidemia | 0.569 | 0.281-1.153 | 0.117 | 0.438 | 0.201-0.953 | **0.038** |
| Hypertension Excluding BMI ≥ 25 | 1.910 | 1.097-3.325 | **0.022** | 1.227 | 0.662-2.273 | 0.516 |
| Diabetes | 1.637 | 0.790-3.395 | 0.185 | 0.754 | 0.326-1.741 | 0.508 |
| Diabetes Excluding Hypertension | 2.721 | 0.977-2.721 | 0.056 | 1.821 | 0.559-5.929 | 0.320 |
| Diabetes Excluding BMI ≥ 25 | 2.086 | 0.882-4.934 | 0.094 | 0.962 | 0.353-2.619 | 0.962 |
| Diabetes Excluding Dyslipidemia | 0.497 | 0.117-2.116 | 0.344 |  |  |  |
| Dyslipidemia | 2.418 | 1.380-4.237 | **0.002** | 2.164 | 1.165-4.021 | **0.015** |
| Dyslipidemia Excluding Hypertension | 1.128 | 0.586-2.151 | 0.726 |  |  |  |
| Dyslipidemia Excluding Diabetes | 1.162 | 0.967-2.858 | 0.066 | 1.915 | 1.039-3.526 | **0.037** |
| Dyslipidemia Excluding BMI ≥ 25 | 2.857 | 1.652-4.924 | **< 0.001** | 2.525 | 1.365-4.669 | **0.003** |
| Dyslipidemia and Hypertension | 2.548 | 1.456-4.458 | **0.001** | 1.721 | 0.927-3.195 | 0.085 |
| Dyslipidemia and Diabetes | 3.329 | 1.432-7.740 | **0.005** | 1.406 | 0.544-3.635 | 0.482 |
| Dyslipidemia and BMI ≥ 25 | 0.826 | 0.379-1.800 | 0.631 | | | |
| BMI $\geq 25$ | 0.522 | 0.275-0.991 | **0.047** | 0.528 | 0.262-1.067 | 0.075 |

Model adjusted for age and sex.

### Analysis of dyslipidemias indexes for GBC

Our results show that dyslipidemia is a risk factor for GBC. We therefore sought to clarify this association with dyslipidemia subtypes; including hypercholesterolemia, TG, increased low density lipoprotein (LDL) and decreased high density lipoproteins (HDL). In both univariate and multivariable analyses, dyslipidemia and decreased HDL were closely associated with increased risk of GBC (Table 5). Combined with the results of Table S2, we found that decreased HDL played an important role in diagnosis or development of GBC. Elevated total cholesterol level and elevated TG level had no significantly impact on GBC status.

**Table 5.**
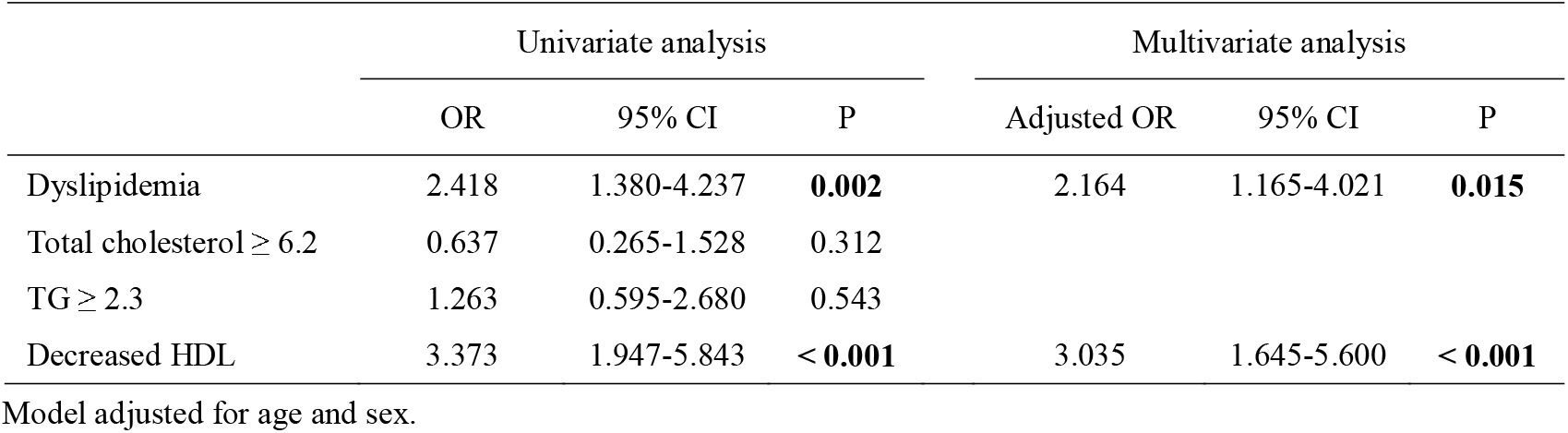
Comparison of different indexes of dyslipidemias for gallbladder lesions

|  | Univariate analysis |  |  | Multivariate analysis |  |  |
| --- | --- | --- | --- | --- | --- | --- |
|  | OR | 95% CI | P | Adjusted OR | 95% CI | P |
| Dyslipidemia | 2.418 | 1.380-4.237 | **0.002** | 2.164 | 1.165-4.021 | **0.015** |
| Total cholesterol $\geq 6.2$ | 0.637 | 0.265-1.528 | 0.312 | | | |
| TG $\geq 2.3$ | 1.263 | 0.595-2.680 | 0.543 | | | |
| Decreased HDL | 3.373 | 1.947-5.843 | **< 0.001** | 3.035 | 1.645-5.600 | **< 0.001** |

Model adjusted for age and sex.

## Discussion

In this study of patients with gallbladder lesions, we elucidated the association between all four metabolic traits – diabetes, hypertension, obese and dyslipidemia – which were individually and jointly associated with others and GBC. We found dyslipidemia was an independent risk factor for GBC, regardless of the presence of diabetes, hypertension and obese. Dyslipidemia was associated with a 2.6-fold increased risk of GBC. Dyslipidemia was present in 44% of patients with gallbladder lesions. This finding suggests that dyslipidemia may contribute to the progression of GBC in a high proportion of patients. As above mentioned, the incidence of GBC is low, but it got poor prognosis[13] and high mortality, and the 5-year survival rate of advanced GBC is very low[14]. There have been some reports of warning factors for GBC that may help diagnose, but unfortunately it is difficult for them to be widely accepted[15] and GBC is therefore frequently diagnosed late. Indeed, most of GBCs are found incidentally by a pathological examination after operation.

Our team believes that based on the results presented, it is still difficult to construct a comprehensive and a widely accepted disease prediction model, so we only discuss the relationship between dyslipidemia and GBC in order to provide new sights for follow-up researches, clinician’s diagnosis and treatment measures, so as to pursue the greatest patients’ benefit. As we know, cholecystectomy is usually performed with gallbladder lesions and a majority of the GBC are discovered incidentally by a pathological examination after operation. So, in order to increase the early diagnosis rate of patients with GBC and further understand the occurrence and development of GBC, we made an overall assessment and analysis of the metabolic characteristics of patients with gallbladder lesions.

We found that age, size of polyps, presence of stones, obesity and dyslipidemia were significantly associated with the GBC group compared to gallbladder lesion group, however this does not mean that these factors are all risk factors for GBC. As previously reported, older age, single polyps, larger polyp diameter, elevated bilirubin level were risk factors of GBC[9, 11, 12], similar to our results. In our multivariate analysis, we found that single polyps and elevated bilirubin level had no significant association, but dyslipidemia was still a significant risk factor for GBC. Patients who are older than 50 years and have the polyps that are larger than 10-mm in diameter have been considered an indication for recommended surgical management[16]. Therefore, we do not discuss in detail in this study, but as a comparative factor to laterally confirm the effect of dyslipidemia on GBC. Interestingly, presence of gallstones appears to be a protective factor for GBC and this point needs to be further verified in our following study.

As diabetes and obesity contribute to the development and progression of numerous malignancies[17], the association between dyslipidemia and neoplasms has also been gradually discovered. Individual metabolic syndrome components are closely related and frequently co-occur. Dyslipidemia is usually a feature in patients with Type 2 diabetes (T2D) and obesity. Patients with obesity and diabetes frequently exhibit elevated circulating cholesterol, TG, small dense low-density lipoprotein (LDL) cholesterol, and reduced HDL cholesterol[18]. Elevated total cholesterol, elevated TG and decreased HDL have been associated with an 18%, 15%, and 20% increased risk of specific cancer, respectively[19]. Besides, previous studies indicated the statin, cholesterol-lowering 3-hydroxy-3-methyl-glutaryl-CoA reductase inhibitors, could decrease the risk of developing specific cancers[20]. Future research to further confirm this relationship would be valuable. Dyslipidemia also has a close relation to oxidative stress. Dyslipidemia downregulates the HDL antioxidant/ anti-inflammatory function which increases oxidative stress and inflammatory, which has been proven to be a contributing factor for specific cancers[21]. Based on the above discussion, we speculate that there could be a correlation between dyslipidemia and the occurrence of GBC.

In addition, there is evidence that dyslipidemia contributes to the formation of benign gallbladder polyps. Bile cholesterol saturation may accelerate cholesterol crystallization, causing subsequent formation of cholesterol polyps [22]. This mechanism could be confirmed in future experiments, as the data generated here cannot confirm this pathway.

The multivariate analysis performed, demonstrated that dyslipidemia is a risk factor related to GBC. However, as discussed above, the relationship between the metabolic syndrome is close, so we further evaluated the interaction between them. Dyslipidemia was associated with a 2.1-fold increase in the odds of GBC. We found that when other metabolic conditions were also present with dyslipidemia, there was no increase the odds of GBC. In contrast, after excluding the interaction of diabetes and obese, the effects of dyslipidemia on GBC were slightly stronger (adjusted OR 1.915 and 2.525, respectively). Previous literatures reported that T2D had an increased risk of GBC both in men and women[23]. The significant statistic difference in diabetes excluding hypertension group maybe due to the less proportion of patients and we would confirm the result furtherly. In healthy people, BMI is typically closely related to blood lipid levels; however, cancers are associated with weight loss and cachexia, hence the BMI of patients cannot accurately reflect the current metabolic state. As our results show, when interference from obesity was excluded, dyslipidemia was associated with a 2.5-fold increase in the odds of GBC having a greater impact than any other metabolic factors. In contrast, obesity seemed to be a protective factor for GBC, so dyslipidemia interplays with obesity had no statistic difference (P=0.631).

HDL is a heterogeneous mixture of spherical macromolecules which varies in size (80-120A)[24]. HDLs possess various functions that have been described including anti-atherosclerotic, anti-oxidative, anti-inflammatory, anti-thrombotic and anti-apoptotic; as well as being involved in immune system modulation and endothelial protection[25]. At present, there are numerous literatures reporting the relation between HDL levels and cancer incidence and mortality, but there is still some discrepancy. Some research discovered the anti-cancer functions of HDLs or HDL-associated molecular components provide a support to the inverse association between HDL level and cancer[26]. Other research reported the association between HDL and the risk of obesity-related cancers, and discovered a modest relation between low HDL level and cancer risk[19]. On the other hand, plasma HDL levels often decrease in patients with cancer, and decreased HDL seems be a typical hallmark of cancer-induced dyslipidemia[27]. However, decreased HDL, similar to dyslipidemia, may be an epiphenomenon of cancer-related inflammation and cancer cell ability which drive cholesterol efflux toward the sites of cancer cell proliferation[28]. So, despite the presence of low HDL level in patients with cancers and association between decreased HDL and cancers, there needs to be more basic research to confirm the impact of hypoxemia on the formation and development of cancers. Low HDL level was associated with an approximately 3-fold increase in the risk of malignancy for patients with gallbladder lesions (Table 5). The gallbladder adenomas are benign neoplastic lesions but are considered with malignant potential. We also identified decreased HDL was associated with a 5-fold increase in the risk of malignancy for patients with neoplastic gallbladder lesions (Table S2). Thus, we suggest patients with gallbladder lesions and low HDL levels should have more frequent follow-ups for early detection of GBC.

This study has some limitations. First, this study is a retrospective analysis of a limited number of enrolled patients. Second, due to the different practices of surgeons and the lack of a unified, standard of diagnosis and treatment, many interesting characteristics including tumor markers, gallbladder wall thickening, patient lifestyles and more were missed. Third, the number of patients in GBC group is small, so there is a moderate risk of selection bias. Finally, no follow-up data for gallbladder lesions patients were available given the retrospective study. The study should furthermore be validated by prospective randomized clinical trials.

In conclusion, our results indicate that dyslipidemia is associated with an increased risk of malignancy in patients with gallbladder lesions, regardless of the presence of hypertension, diabetes and obesity. We suggest patients with gallbladder lesions and low HDL levels should have more frequent follow-ups for early detection of GBC.

## Data Availability

All data are available

## Abbreviations

BGP: Benign gallbladder polyps
GBC: Gallbladder cancer
TG: Triglycerides
HDL: High-density lipoprotein
BMI: body mass index
OR: odds ratio
CI: confidence interval
HDL: high density lipoprotein

## Acknowledgements

This study was funded by Cultivating Funds of Renji Hospital, School of Medicine, Shanghai Jiao Tong University, PYZY16-017.

## Author contributions

Zheng Deng, Yan Xuan and Xinxing Li contributed equally to this work. Study design: Haolu Wang, Tao Chen; Data collection: Zheng Deng, Yan Xuan, Xinxing Li, Zhiqing Yuan and Zhoukan Chen; Statistical analysis: Zheng Deng and Haolu Wang; Manuscript: Zheng Deng, William J Crawford and Anastasia Brooks. Critical revision of the manuscript: Yan Xuan, Xinxing Li, Xiaowen Liang and Tao Chen.

## Compliance with ethical standards

### Ethical approval

This study was conducted in accordance with the Declaration of Helsinki. The study protocol was approved by the Shanghai Jiaotong University School of Medicine, Renji Hospital Ethics Committee (Ethical Approval Nomber 2016-045).

